# Validity of denture usage definitions based on claims data in Japanese older adults: The Longevity Improvement and Fair Evidence (LIFE) Study

**DOI:** 10.1101/2025.10.17.25338212

**Authors:** Anna Kinugawa, Yudai Tamada, Taro Kusama, Manami Hoshi-Harada, Sachiko Ono, Futoshi Oda, Megumi Maeda, Nobuhiro Yoda, Ken Osaka, Haruhisa Fukuda, Kenji Takeuchi

## Abstract

**Background:** Administrative claims data are increasingly used in oral health research, however, their validity for identifying denture use has not been established.

**Objective:** To evaluate the accuracy of algorithms based on dental claims codes for identifying denture users, dentist-reported oral health screening records were used as the reference standard.

**Methods:** We analyzed data from 4,053 adults aged ≥65 years in the Longevity Improvement and Fair Evidence (LIFE) Study. Twelve algorithms incorporating 56 denture-related disease and procedure codes were developed from claims in the 12 months preceding the screening. Sensitivity, specificity, positive predictive value (PPV), and negative predictive value (NPV) were calculated. Sensitivity analyses used a 6-month claims window. A post hoc bias analysis was also performed.

**Results:** During screening, 59.9% of the participants were classified as denture users. Algorithms based solely on disease codes showed very high specificity (98.3–100%) but low sensitivity (1.4– 20.8%). Among the procedure-based definitions, oral rehabilitation (algorithm f) had the highest sensitivity (63.6%). The combined algorithm using both disease and procedure codes (algorithm C) achieved the best balance, with 65.3% sensitivity, 96.6% specificity, 96.6% PPV, and 65.1% NPV. Similar findings were observed using the 6-month claims data. Bias analysis indicated that the risks for denture use could be underestimated by 26–59%, with algorithm C showing the least bias.

**Conclusions:** Denture use can be identified from dental claims data with moderate accuracy. Algorithms combining denture-related disease and procedure codes, or using oral rehabilitation codes, provide practical definitions for research on oral and systemic health outcomes.

## 1. Backgrounds

Administrative claims data have been increasingly used in epidemiological and health service research in recent years because they provide large sample sizes, population representativeness, and insights into real-world effectiveness and clinical practice patterns^1^. However, because claims are generated for fee-for-service reimbursements rather than for research purposes, data inaccuracies may lead to misclassification and bias^2,3^. Therefore, validation against a reference standard is therefore essential.

The use of dentures may mitigate the risk of various systemic health outcomes that increase with tooth loss. Previous studies have shown that compared with non-denture users, denture users have a lower risk of various outcomes associated with tooth loss, indicating physical or cognitive health, such as mortality, need for long-term care, and dementia^4^, psychological health, such as depressive symptoms^5^ and low frequency of laughter^6^, and health behaviors, such as reduced protein intake^7^. Denture use has also been reported to moderate the risk of incident pneumonia associated with dysphagia^8^. In light of these findings, it has recently been suggested that denture use may also contribute to the maintenance of quality of life in older adults^9^.

Previous validation studies have demonstrated that although the number of remaining teeth can be ascertained using dental claims data^10-12^, whether denture use can be accurately identified has not yet been evaluated. If denture use could be reliably measured through administrative claims data, this would substantially expand the opportunities for oral health research. For example, it would enable large-scale studies on the role of prosthetic rehabilitation in aging populations, facilitate health service evaluations of access to and equity in denture provision, and support investigations on the impact of denture use on systemic health outcomes and long-term care needs.

The present study aimed to examine the validity of claims-based algorithms in identifying denture use in Japanese older adults. We compared claims-based definitions based on denture-related disease and procedure codes with dentist-assessed denture usage from oral healthcare screening records, which served as the reference standard.

## 2. Methods

### 2.1. Setting

This validation study used the Longevity Improvement and Fair Evidence (LIFE) Study database from one Japanese municipality, covering the period from April 2018 to March 2020, which includes two health insurance (the National Health Insurance (NHI) and the Latter-Stage Older Persons Health Care System) claims data and oral healthcare screening data^13^.

In the Japanese health insurance system, NHI covers the self-employed or not employed, including part-time workers and retired individuals, and their dependents aged <75 years; the Later-Stage Older Persons Health Care System covers individuals aged ≥75 years. The details of the Japanese health insurance system are described elsewhere^14^. This health insurance system covers conventional dentures such as acrylic resin dentures, excluding metal plate dentures and implant-supported dentures^15^. The present validity study targeted individuals wearing conventional dentures.

In the target municipality, oral healthcare screening is provided free of charge to residents aged 65 years and older to promote their oral health and prevent the need for long-term care. The program was implemented in 2018 and had a participation rate of approximately 5%.

The target population included individuals who underwent oral healthcare screening in the fiscal year 2019, had at least one dental claim in the 12 months prior to the screening month, and were continuously enrolled in either the NHI or the Latter-Stage Older Persons Health Care System during the study period (April 2018 to March 2020). The following records were excluded: non-assigned ID, duplicate records, and incorrect dates. A flowchart of the study participants is shown in Figure 1.

**Figure 1.**
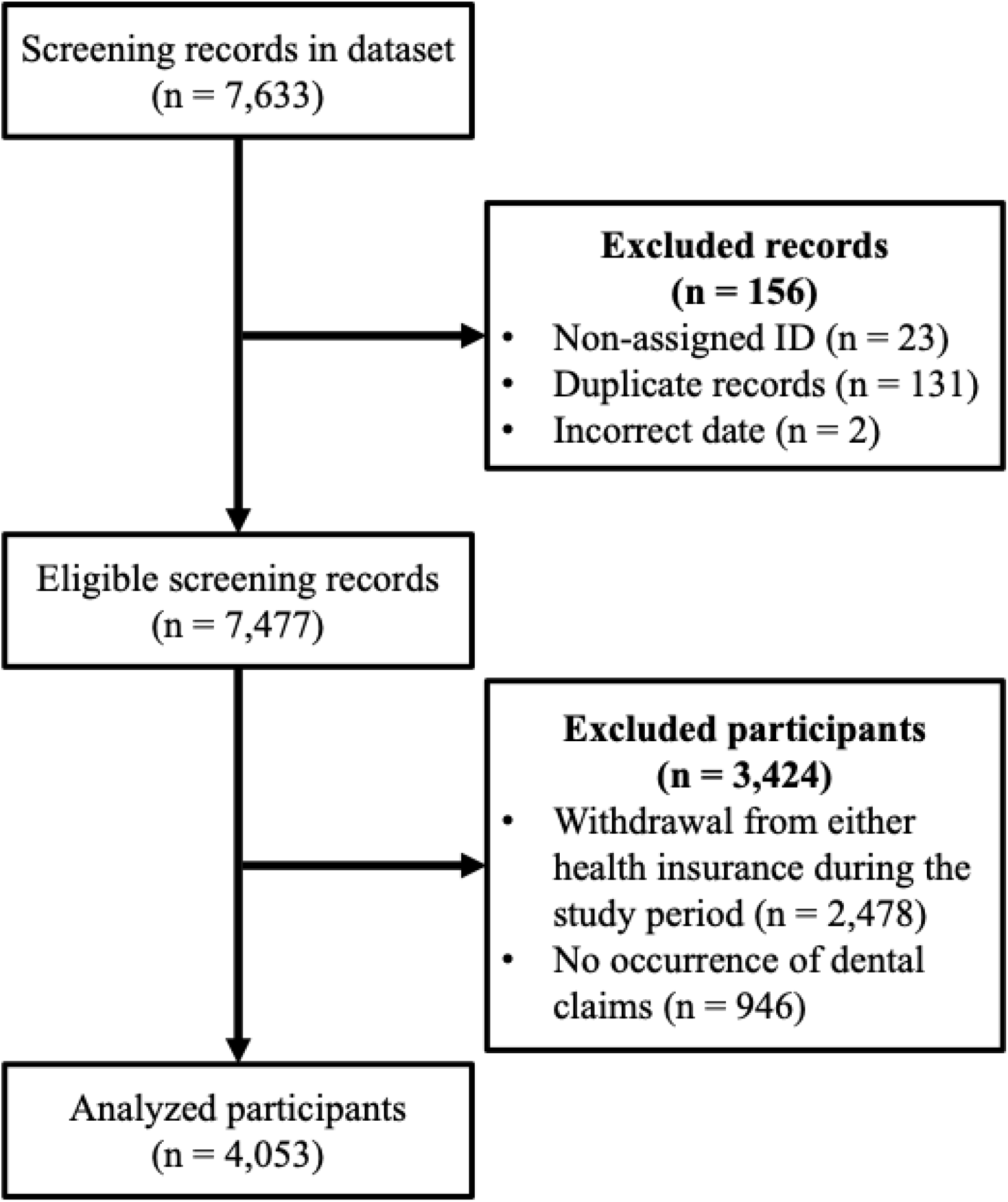
Flowchart of analyzed participants.

### 2.2. Reference standard and algorithms

The record of using or not using dentures, based on an assessment by a dentist during oral healthcare screening, was used as the reference standard.

We used 12 algorithms with 56 denture-related disease and procedure codes in the 12 months of dental claims data prior to the screening month (Supplementary Table 1). Supplementary Table 2 shows the distribution of the disease and procedure code records. These algorithms comprised four disease records (a: ill-fitting denture, b: broken denture, c: missing teeth, d: other denture-related diseases), five procedure records (e: management of new dentures, f: oral rehabilitation, g: denture repair, h: denture relining, i: other denture-related procedures), and three combinations of disease and procedure records (A: denture-related diseases (a–d), B: all denture-related procedures (e–i), and C: either denture-related diseases or procedures (A or B)).

### 2.3. Statistical analysis

Sensitivity, specificity, positive predictive value (PPV), and negative predictive value (NPV) with 95% confidence intervals (95% CIs) were calculated for each algorithm. Sensitivity and specificity were defined as the proportions of true denture users and non-users, respectively, correctly identified by each algorithm. The PPV was defined as the proportion of individuals identified as denture users based on claims that were confirmed as users at screening. The NPV was defined as the proportion of individuals identified as non-users based on claims that were confirmed as non-users during screening. For sensitivity analysis, we defined the algorithms using the participants’ 6 months dental claims data prior to the screening month.

To estimate the potential impact of misclassification of exposure when denture use was applied as an exposure variable in epidemiological studies, we performed a post-hoc bias estimation analysis for algorithms A–C^16^. Percent bias, defined as the difference between the observed relative risk (RR_O_) and the true relative risk (RR_T_) divided by the RR_T_ using the sensitivity and specificity obtained from the present study, was simulated by setting the RR_T_ to 2.00 and 3.00, and by using the prevalence of denture use determined based on the results from this screening and the 2022 Survey of Dental Disease^17^.

All analyses were performed using STATA software (version 17.0; Stata Corp., College Station, TX, USA). This study was conducted according to the checklist of reporting criteria for studies validating health administrative data algorithms^18^.

### 2.4 Ethical approval

This study was approved by the Kyushu University Institutional Review Board for Clinical Research (approval No. 226530) and the Ethics Committee of Tohoku University Graduate School of Dentistry (approval No. 40589). Approval for data use was obtained from the municipality’s Personal Information Protection Review Board.

## 3. Results

Overall, 4,053 participants were included in the present study, and 2,427 participants (59.9%) were defined as denture users based on oral healthcare screening. The participants’ characteristics are listed in Table 1. The mean age of the participants was 78.2 years (SD = 6.3), and 61.4% were women. Denture users had fewer teeth than non-users (mean: 13.7 vs. 25.0).

**Table 1.** Characteristics of study participants (n = 4,053).

|  | Non-denture users<br>(n = 1,626) | Denture users<br>(n = 2,427) | Total<br>(n = 4,053) |
| --- | --- | --- | --- |
| Age [n, (%)] |  |  |  |
| 65–74 | 509 (55.4) | 410 (44.6) | 919 (22.7) |
| ≥75 | 1,117 (35.6) | 2,017 (64.4) | 3,134 (77.3) |
| Sex [n, (%)] |  |  |  |
| Men | 618 (39.5) | 948 (60.5) | 1,566 (38.6) |
| Women | 1,008 (40.5) | 1,479 (59.5) | 2,487 (61.4) |
| Number of teeth [mean, (SD)] | 25.0 (3.7) | 13.7 (7.7) | 18.2 (8.5) |

Abbreviation: SD, standard deviation.

Table 2 shows the number of participants with true positives (TP), false negatives (FN), false positives (FP), true negatives (TN), and the accuracy of the algorithms in identifying denture users using dental claims data from 12 months prior to oral healthcare screening. In the algorithms using denture-related disease codes (a–d), the sensitivity ranged from 1.4% (b: broken denture) to 20.8% (a: ill-fitting denture). The specificity and NPV ranged from 98.3% (a) to 100% (b–d) and from 40.5% (b–d) to 45.4% (a), respectively. The PPV ranged from 94.7% (a) to 100% (d: other denture-related diseases). Among the algorithms using denture-related procedure codes (e–i), algorithm f (oral rehabilitation) demonstrated the highest sensitivity (63.6%) and NPV (64.0%), but its specificity (96.7%) and PPV (96.7%) were slightly lower than those of the other procedure-based algorithms. Among the combined algorithms, algorithm C (any denture-related disease code or procedure code) achieved the highest sensitivity (65.3%) and NPV (65.1%) along with moderate specificity (96.6%) and PPV (96.6%). Its overall performance was comparable to that of algorithm B using all denture-related procedure codes (sensitivity, 64.3%; specificity, 96.7%; PPV, 96.7%; and NPV, 64.5%).

**Table 2.**
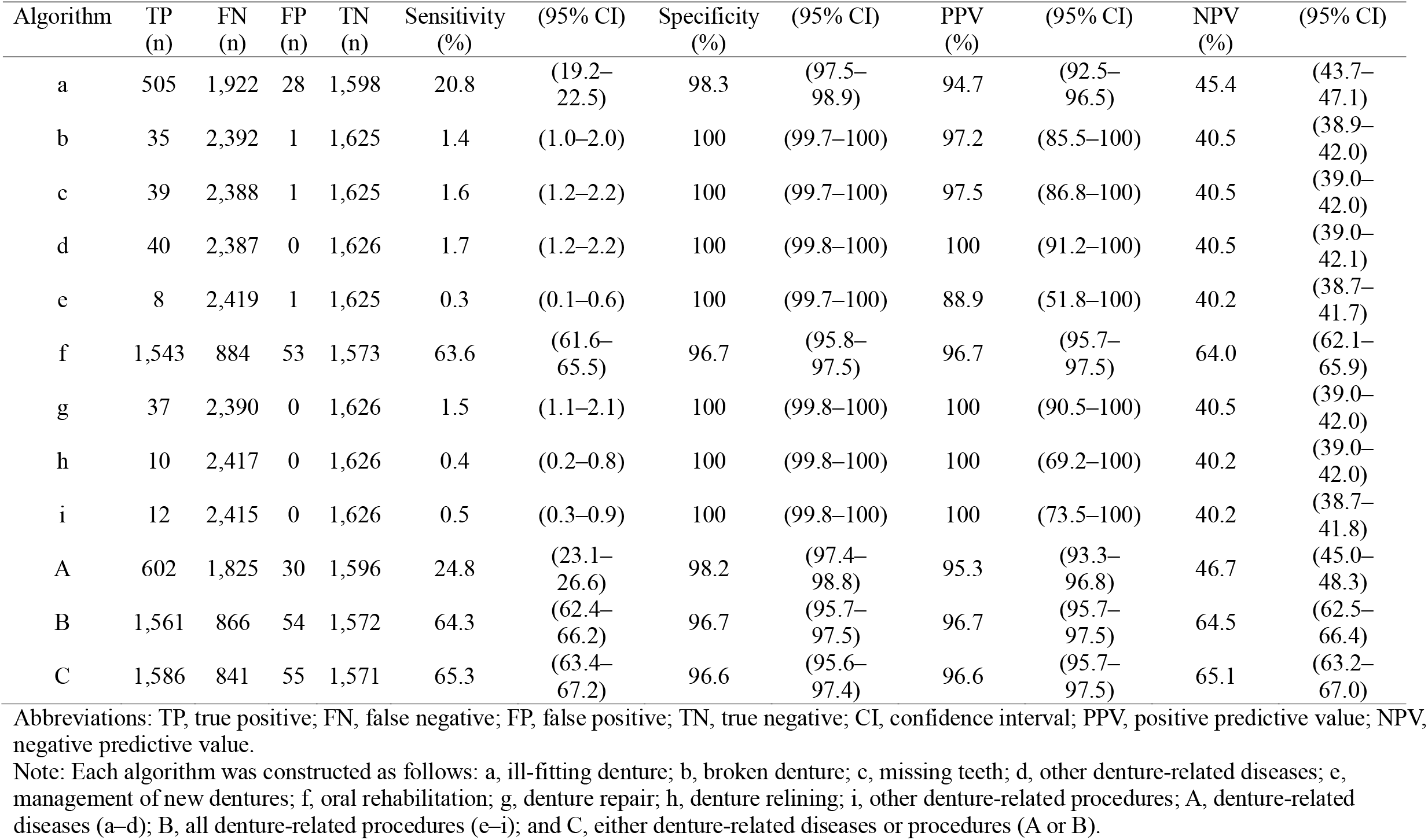
Accuracy of algorithms in identifying denture users from 12-month claims data before oral healthcare screening (n = 4,053).

In the sensitivity analysis using 6 months dental claims data prior to the screening month, similar estimates of accuracy were observed, as in the main result of 12 months analysis (Supplementary Table 3).

The results of the post-hoc bias estimation analysis using the sensitivity and specificity of algorithms A–C from the present study are presented in Supplementary Table 4. In the case where the exposure of denture usage was determined based on the results from this screening (the proportion exposed to X was set to 0.599 in scenarios 1 and 2), the percent bias in RR ranged from - 36.1 to -26.4 when the RR_T_ was 2.00, and -52.9 to -41.5 when the RR_T_ was 3.00. If the exposure of denture usage was determined based on the 2022 Survey of Dental Disease (the proportion exposed to X was set to 0.748 in scenarios 3 and 4), the percent bias in RR ranged from -41.5 to -33.9 when the RR_T_ was 2.00, and -58.6 to -50.6 when the RR_T_ was 3.00. Algorithm C yielded the smallest bias across the scenarios.

## 4. Discussion

This validation study examined the accuracy of algorithms for identifying denture users from Japanese dental claims data using dentist-reported screening records as the reference standard. Among the algorithms tested, the combination of denture-related disease and procedure codes (algorithm C) achieved the highest sensitivity (65.3%) and NPV (65.1%), with moderate specificity (96.6%) and PPV (96.6%). This approach demonstrated greater agreement with the reference standard than disease- or procedure-based algorithms alone.

Notably, algorithm f (oral rehabilitation) also showed strong performance, with relatively high sensitivity (63.6%) and accuracy across all measures. Together with algorithms B and C, this represents a practical approach for identifying denture users from claims data, particularly in studies where sensitivity is prioritized.

Although some algorithms (e.g., broken denture or denture repair) exhibited very high specificity and PPV, they were limited by their low sensitivity because they capture only a small subset of denture users. By contrast, algorithms C, B, and f performed consistently across metrics, suggesting that broader code sets or specific procedure codes with high coverage are best suited for identifying denture use in claims data.

The sensitivity analysis, which used 6 months of dental claims data before the screening month, yielded results broadly consistent with those of the main analysis, which used 12 months of data. This suggests that denture use can be identified within a relatively short observation period. However, previous studies have reported that most denture users do not visit the dentist regularly for denture maintenance every 6 months or annually^19,20^. Therefore, a longer follow-up period may be more appropriate for research contexts in which maximizing case ascertainment is critical. An optimal observation window should be selected based on the objectives of the study.

The post-hoc bias analysis highlighted the implications of misclassification when denture use was applied as an exposure variable in epidemiologic research. Across scenarios, the algorithms underestimated the true RRs by 26–59%, with algorithm C producing the smallest bias. These findings underscore the importance of accounting for exposure misclassification when interpreting the results based on claim-derived denture data.

This study had some limitations. First, participants were restricted to a single municipality, and individuals were continuously enrolled in the NHI or the Latter-Stage Older Persons Health Care System. Exclusions owing to age-related transitions in insurance coverage^14^ may have introduced a selection bias. Second, the algorithms were validated within the Japanese claims system, which may differ in coding practices from other countries, limiting generalizability.

Therefore, external validation in other settings is required. Despite these limitations, this study provides important evidence of the feasibility of using claims data to identify denture use. In combination with information on the number of remaining teeth, such measures can expand opportunities to assess oral health status and its relationship with systemic health outcomes in large-scale epidemiologic studies.

## 5. Conclusion

This validation study demonstrated that denture use can be identified from dental claims data with moderate accuracy. The combined algorithm using both denture-related disease and procedure codes (algorithm C) achieved the highest sensitivity and NPV while maintaining high specificity and PPV. Algorithm f (oral rehabilitation) also showed high accuracy, indicating its potential utility in claims-based studies. Although misclassification remains a concern, particularly for exposure assessments in epidemiological research, these algorithms provide a practical approach for incorporating denture use in the analyses of oral systemic health.

## Acknowledgements

We are grateful to other investigators, the staff, and the participants for LIFE Study for their valuable contributions. AK was supported by the MEXT/JSPS WISE Program: Advanced Graduate Program for Future Medicine and Health Care of Tohoku University and JST SPRING (JPMJSP2114).

## Abbreviations

PPV: positive predictive value;
NPV: negative predictive value;
NHI: National Health Insurance;
95%CIs: 95% confidence intervals;
RR: relative risk;
TP: true positive;
FN: false negative;
FP: false positive;
TN: true negative;
SD: standard deviation.

## Supplementary material

Supplemental material is available online.

## Data availability statement

All datasets used in this study are not publicly available to ethical or legal restrictions. For inquiries regarding the datasets used in this study, please contact Dr. Haruhisa Fukuda, Principal Investigator of the LIFE Study, with reasonable requests.

