## Supplementary material for "Validity of denture usage definitions based on claims data in Japanese older adults: The Longevity Improvement and Fair Evidence (LIFE) Study"

**Supplementary Table 1.** List of using Disease/Procedure codes.

| Disease/Procedure code No. | Disease/Procedure code name (EN) | Disease/Procedure code name (JP) | Procedure/Disease code |
|---|---|---|---|
| **Disease** | | | |
| **Algorithm a: Ill-fitting denture** | | | |
| 1 | Ill-fitting denture | 義歯不適合 | 8842706 |
| 2 | Ill-fitting denture base | 義歯床不適合 | 8848868 |
| 3 | Ill-fitting mucosal surface of denture base | 義歯床粘膜面不適合 | 8843863 |
| 4 | Ill-fitting occlusal surface of denture | 義歯咬合面不適合 | 9999527 |
| 5 | Ill-fitting partial denture | 局部義歯不適合 | 8845550 |
| 6 | Ill-fitting mucosal surface of partial denture base | 局部義歯義歯床粘膜面不適合 | 8845545 |
| 7 | Ill-fitting occlusal surface of partial denture | 局部義歯咬合面不適合 | 8845546 |
| 8 | Ill-fitting complete denture | 総義歯不適合 | 8845580 |
| 9 | Ill-fitting mucosal surface of complete denture base | 総義歯義歯床粘膜面不適合 | 8845575 |
| 10 | Ill-fitting occlusal surface of complete denture | 総義歯咬合面不適合 | 8845576 |
| 11 | Ill-fitting old denture | 旧義歯不適合 | 8846555 |
| **Algorithm b: Broken denture** | | | |
| 12 | Broken denture | 義歯破損 | 9999524 |
| 13 | Broken partial denture | 局部義歯破損 | 8845549 |
| 14 | Broken complete denture | 総義歯破損 | 8845579 |
| 15 | Broken old denture | 旧義歯破損 | 8845646 |
| **Algorithm c: Missing teeth** | | | |
| 16 | Missing teeth | 欠損歯 | 5250001 |
| 17 | Missing teeth/base fitting | 欠損歯・床適合 | 8845780 |
| 18 | Missing teeth/add tooth | 欠損歯・増歯 | 8844171 |
| 19 | Missing teeth/reline denture | 欠損歯・裏装 | 8844173 |
| **Algorithm d: Other denture-related diseases** | | | |
| 20 | Denture fibroma | 義歯性線維腫 | 5238007 |
| 21 | Denture stomatitis | 義歯性口内炎 | 8832750 |
| 22 | Oral decubitus ulcer | 口腔褥瘡性潰瘍 | 8844867 |
| 23 | Denture-related ulcer | 義歯性潰瘍 | 8845551 |
| 24 | Mucosal abnormalities under denture base | 義歯床下粘膜異常 | 8843691 |
| 25 | Mucosal abnormalities under partial denture base | 局部義歯義歯床下粘膜異常 | 8845547 |
| 26 | Mucosal abnormalities under complete denture base | 総義歯義歯床下粘膜異常 | 8845577 |
| 27 | Denture impaction | 義歯嵌入 | 8842373 |
| 28 | Denture overhanging | 義歯過高 | 8843862 |
| 29 | Denture underhanging | 義歯低位 | 8843864 |
| 30 | Partial denture impaction | 局部義歯嵌入 | 8845544 |
| 31 | Partial denture overhanging | 局部義歯過高 | 8845543 |
| 32 | Partial denture underhanging | 局部義歯低位 | 8845548 |
| 33 | Complete denture overhanging | 総義歯過高 | 8845574 |

| | | | |
|---|---|---|---|
| 34 | Complete denture underhanging | 総義歯低位 | 8845578 |
| **Procedure** | | | |
| **Algorithm e: Management of new dentures** | | | |
| 35 | Management of new denture (per oral cavity) | 新製有床義歯管理料（１口腔につき）（２以外の場合） | 302003710 |
| 36 | Management of new denture (per oral cavity) (in case of difficulty) | 新製有床義歯管理料（１口腔につき）（困難な場合） | 302008310 |
| **Algorithm f: Oral rehabilitation** | | | |
| 37 | Oral rehabilitation (per oral cavity) | 歯科口腔リハビリテーション料１（有床義歯の口以外の場合）（１口腔につき） | 308002510 |
| 38 | Oral rehabilitation (in case of difficulty) (per oral cavity) | 歯科口腔リハビリテーション料１（有床義歯の困難な場合）（１口腔につき） | 308002610 |
| **Algorithm g: Denture repair** | | | |
| 39 | Denture repair (per denture) | 有床義歯修理（１床につき） | 313021610 |
| 40 | Denture repair (per denture) (within 6 months of initial delivery) | 有床義歯修理（１床につき）（６月以内） | 313021770 |
| **Algorithm h: Denture relining** | | | |
| 41 | Denture relining using hard material (partial denture, per denture, 1–4 teeth) | 有床義歯内面適合法（硬質材料を用いる場合（局部義歯（１床につき）（１歯から４歯まで））） | 313021810 |
| 42 | Denture relining using hard material (partial denture, per denture, 5–8 teeth) | 有床義歯内面適合法（硬質材料を用いる場合（局部義歯（１床につき）（５歯から８歯まで））） | 313021910 |
| 43 | Denture relining using hard material (partial denture, per denture, 9–11 teeth) | 有床義歯内面適合法（硬質材料を用いる場合（局部義歯（１床につき）（９歯から１１歯まで））） | 313022010 |
| 44 | Denture relining using hard material (partial denture, per denture, 12–14 teeth) | 有床義歯内面適合法（硬質材料を用いる場合（局部義歯（１床につき）（１２歯から１４歯まで））） | 313022110 |
| 45 | Denture relining using hard material (complete denture, per jaw) | 有床義歯内面適合法（硬質材料を用いる場合（総義歯（１顎につき）） | 313022210 |
| 46 | Denture relining using soft material (complete denture, per jaw) | 有床義歯内面適合法（軟質材料を用いる場合（１顎につき）） | 313028610 |

| | | | |
|---|---|---|---|
| 47 | Denture relining (within 6 months of initial delivery) | 有床義歯内面適合法（６月以内） | 313028770 |
| **Algorithm i: Other denture-related procedures** | | | |
| 48 | Tissue conditioning under denture base (per jaw) | 有床義歯床下粘膜調整処置（１顎１回につき） | 309008310 |
| 49 | Masticatory function test with denture (per oral cavity) (in case of measurement of lower jaw movement and masticatory ability) | 有床義歯咀嚼機能検査（１口腔につき）（有床義歯咀嚼機能検査１（１回につき）（下顎運動測定と咀嚼能力測定を併せて行う場合）） | 304001910 |
| 50 | Masticatory function test with denture (per oral cavity) (in case of measurement of masticatory ability) | 有床義歯咀嚼機能検査（１口腔につき）（有床義歯咀嚼機能検査１（１回につき）（咀嚼能力測定のみを行う場合）） | 304002010 |
| 51 | Masticatory function test with denture (per oral cavity) (in case of measurement of lower jaw movement and occlusal pressure) | 有床義歯咀嚼機能検査（１口腔につき）（有床義歯咀嚼機能検査２（１回につき）（下顎運動測定と咬合圧測定を併せて行う場合）） | 304002210 |
| 52 | Masticatory function test with denture (per oral cavity) (in case of measurement of occlusal pressure) | 有床義歯咀嚼機能検査（１口腔につき）（有床義歯咀嚼機能検査２（１回につき）（咬合圧測定のみを行う場合）） | 304002310 |
| 53 | Additional payment for cooperation with dental technician 1 (per denture) (for denture repair) | 歯科技工加算１（１床につき）（有床義歯修理） | 313028570 |
| 54 | Additional payment for cooperation with dental technician 2 (per denture) (for denture relining) | 歯科技工加算２（１床につき）（有床義歯修理） | 313023670 |
| 55 | Additional payment for cooperation with dental technician 1 (per jaw) (for denture repair) | 歯科技工加算１（１顎につき）（有床義歯内面適合法） | 313030770 |
| 56 | Additional payment for cooperation with dental technician 2 (per jaw) (for denture relining) | 歯科技工加算２（１顎につき）（有床義歯内面適合法） | 313030870 |

Abbreviations: EN, English; JP, Japanese.

**Supplementary Table 2.** Distribution of Disease/Procedure code records.

| Disease/Procedure code No. | No record | Recorded |
|---|---|---|
| 1 | 3,565 (88.0) | 488 (12.0) |
| 2 | 4,043 (99.8) | 10 (0.2) |
| 3 | 4,053 (100) | – |
| 4 | 4,053 (100) | – |
| 5 | 4,025 (99.3) | 28 (0.7) |
| 6 | 4,053 (100) | – |
| 7 | 4,053 (100) | – |
| 8 | 4,046 (99.8) | 7 (0.2) |
| 9 | 4,053 (100) | – |
| 10 | 4,053 (100) | – |
| 11 | 4,053 (100) | – |
| 12 | 4,018 (99.1) | 35 (0.9) |
| 13 | 4,053 (100) | – |
| 14 | 4,052 (100) | 1 (0.0) |
| 15 | 4,053 (100) | – |
| 16 | 4,018 (99.1) | 35 (0.9) |
| 17 | 4,053 (100) | – |
| 18 | 4,050 (99.9) | 3 (0.1) |
| 19 | 4,051 (100) | 2 (0.0) |
| 20 | 4,053 (100) | – |
| 21 | 4,053 (100) | – |
| 22 | 4,023 (99.3) | 30 (0.7) |
| 23 | 4,053 (100) | – |
| 24 | 4,043 (99.8) | 10 (0.2) |
| 25 | 4,053 (100) | – |
| 26 | 4,053 (100) | – |
| 27 | 4,053 (100) | – |
| 28 | 4,053 (100) | – |
| 29 | 4,053 (100) | – |
| 30 | 4,053 (100) | – |
| 31 | 4,053 (100) | – |
| 32 | 4,053 (100) | – |
| 33 | 4,053 (100) | – |
| 34 | 4,053 (100) | – |
| 35 | 4,050 (99.9) | 3 (0.1) |
| 36 | 4,049 (99.9) | 4 (0.1) |
| 37 | 3,181 (78.5) | 872 (21.5) |
| 38 | 3,328 (82.1) | 725 (17.9) |
| 39 | 4,016 (99.1) | 37 (0.9) |
| 40 | 4,053 (100) | – |
| 41 | 4,053 (100) | – |
| 42 | 4,053 (100) | – |
| 43 | 4,052 (100) | 1 (0.0) |
| 44 | 4,052 (100) | 1 (0.0) |
| 45 | 4,050 (99.9) | 3 (0.1) |
| 46 | 4,053 (100) | – |
| 47 | 4,053 (100) | – |
| 48 | 4,036 (99.6) | 17 (0.4) |
| 49 | 4,053 (100) | – |
| 50 | 4,053 (100) | – |
| 51 | 4,053 (100) | – |
| 52 | 4,053 (100) | – |
| 53 | 4,053 (100) | – |
| 54 | 4,053 (100) | – |
| 55 | 4,053 (100) | – |
| 56 | 4,053 (100) | – |

**Supplementary Table 3.** Accuracy of algorithms in identifying denture users from 6-month claims data before oral healthcare screening (n = 3,618).

| Algorithm | TP (n) | FN (n) | FP (n) | TN (n) | Sensitivity (%) | (95% CI) | Specificity (%) | (95% CI) | PPV (%) | (95% CI) | NPV (%) | (95% CI) |
|---|---|---|---|---|---|---|---|---|---|---|---|---|
| a | 519 | 1,674 | 35 | 1,390 | 23.7 | (21.9–25.5) | 97.5 | (96.6–98.3) | 93.7 | (91.3–95.6) | 45.4 | (43.6–47.1) |
| b | 39 | 2,154 | 0 | 1,425 | 1.8 | (1.3–2.4) | 100 | (99.7–100) | 100 | (91.0–100) | 39.8 | (38.2–41.4) |
| c | 57 | 2,136 | 0 | 1,425 | 2.6 | (2.0–3.4) | 100 | (99.7–100) | 100 | (93.7–100) | 40.0 | (38.4–41.6) |
| d | 37 | 2,156 | 1 | 1,424 | 1.7 | (1.2–2.3) | 100 | (99.6–100) | 97.4 | (86.2–100) | 39.8 | (38.2–41.4) |
| e | 3 | 2,190 | 0 | 1,425 | 0.1 | (0.0–0.4) | 100 | (99.7–100) | 100 | (29.2–100) | 39.4 | (37.8–41.0) |
| f | 1,453 | 740 | 56 | 1,369 | 66.3 | (64.2–68.2) | 96.1 | (94.9–97.0) | 96.3 | (95.2–97.2) | 64.9 | (62.8–67.0) |
| g | 33 | 2,160 | 0 | 1,425 | 1.5 | (1.0–2.1) | 100 | (99.7–100) | 100 | (89.4–100) | 39.7 | (38.1–41.4) |
| h | 12 | 2,181 | 0 | 1,425 | 0.5 | (0.3–1.0) | 100 | (99.7–100) | 100 | (73.5–100) | 39.5 | (38.1–41.4) |
| i | 11 | 2,182 | 1 | 1,424 | 0.5 | (0.3–0.9) | 100 | (99.6–100) | 91.7 | (61.5–100) | 39.5 | (37.9–41.1) |
| A | 631 | 1,562 | 35 | 1,390 | 28.8 | (26.9–30.7) | 97.5 | (96.6–98.3) | 94.7 | (92.8–96.3) | 47.1 | (45.3–48.9) |
| B | 1,459 | 734 | 57 | 1,368 | 66.5 | (64.5–68.5) | 96.0 | (94.8–97.0) | 96.2 | (95.2–97.1) | 65.1 | (63.0–67.1) |
| C | 1,489 | 704 | 57 | 1,368 | 67.9 | (65.9–69.8) | 96.0 | (94.8–97.0) | 96.3 | (95.2–97.2) | 66.0 | (63.9–68.1) |

Abbreviations: TP, true positive; FN, false negative; FP, false positive; TN, true negative; CI, confidence interval; PPV, positive predictive value; NPV, negative predictive value.

Note: Each algorithm was constructed as follows: a, ill-fitting denture; b, broken denture; c, missing teeth; d, other denture-related diseases; e, management of new dentures; f, oral rehabilitation; g, denture repair; h, denture relining; i, other denture-related procedures; A, denture-related diseases (a–d); B, all denture-related procedures (e–i); and C, either denture-related diseases or procedures (A or B).

**Supplementary Table 4.** Results from post-hoc bias estimation analysis for algorithms A to C.

| Scenario | Proportion exposed to X† | Incidence of Y in unexposed | Incidence of Y in exposed | Incidence of Y overall | True relative risk | Algorithm sensitivity for X | Algorithm specificity for X | Observed relative risk | **% bias in RR (%)** |
|---|---|---|---|---|---|---|---|---|---|
| | P | $I_U$ | $I_E$ | | $RR_T$ | sens | spec | $RR_O$ | |
| 1 | | | | | | | | | |
| A | 0.599 | 0.01 | 0.02 | 0.02 | 2.00 | 0.288 | 0.975 | 1.28 | **-36.1** |
| B | 0.599 | 0.01 | 0.02 | 0.02 | 2.00 | 0.665 | 0.960 | 1.46 | **-27.0** |
| C | 0.599 | 0.01 | 0.02 | 0.02 | 2.00 | 0.679 | 0.960 | 1.47 | **-26.4** |
| 2 | | | | | | | | | |
| A | 0.599 | 0.03 | 0.09 | 0.07 | 3.00 | 0.288 | 0.975 | 1.41 | **-52.9** |
| B | 0.599 | 0.03 | 0.09 | 0.07 | 3.00 | 0.665 | 0.960 | 1.73 | **-42.2** |
| C | 0.599 | 0.03 | 0.09 | 0.07 | 3.00 | 0.679 | 0.960 | 1.75 | **-41.5** |
| 3 | | | | | | | | | |
| A | 0.748 | 0.01 | 0.02 | 0.02 | 2.00 | 0.288 | 0.975 | 1.17 | **-41.5** |
| B | 0.748 | 0.01 | 0.02 | 0.02 | 2.00 | 0.665 | 0.960 | 1.31 | **-34.4** |
| C | 0.748 | 0.01 | 0.02 | 0.02 | 2.00 | 0.679 | 0.960 | 1.32 | **-33.9** |
| 4 | | | | | | | | | |
| A | 0.748 | 0.03 | 0.09 | 0.07 | 3.00 | 0.288 | 0.975 | 1.24 | **-58.6** |
| B | 0.748 | 0.03 | 0.09 | 0.07 | 3.00 | 0.665 | 0.960 | 1.47 | **-51.1** |
| C | 0.748 | 0.03 | 0.09 | 0.07 | 3.00 | 0.679 | 0.960 | 1.48 | **-50.6** |

Note: Note: Each algorithm was constructed as follows: A, denture-related diseases; B, all denture-related procedures; C, either of denture-related diseases or procedures.

†The proportion exposed to X was based on the results from this screening in scenarios 1 and 2, and on the 2022 Survey of Dental Disease in scenarios 3 and 4.
